# Development of mass spectrometry-based targeted assay for direct detection of novel SARS-CoV-2 coronavirus from clinical specimens

**DOI:** 10.1101/2020.08.05.20168948

**Authors:** Santosh Renuse, Patrick M. Vanderboom, Anthony D. Maus, Jennifer V. Kemp, Kari M. Gurtner, Anil K. Madugundu, Sandip Chavan, Jane A. Peterson, Benjamin J. Madden, Kiran K. Mangalaparthi, Dong-Gi Mun, Smrita Singh, Benjamin R. Kipp, Surendra Dasari, Ravinder J. Singh, Stefan K. Grebe, Akhilesh Pandey

## Abstract

The COVID-19 pandemic caused by severe acute respiratory syndrome-coronavirus 2 (SARS-CoV-2) has overwhelmed health systems worldwide and highlighted limitations of diagnostic testing. Several types of diagnostics including RT-PCR-based assays, antigen detection by lateral flow assays and antibody-based assays have been developed and deployed in a short time. However, many of these assays are lacking in sensitivity and/or specificity. Here, we describe an immunoaffinity purification followed by high resolution mass spectrometry-based targeted assay capable of detecting viral antigen in nasopharyngeal swab samples of SARS-CoV-2 infected individuals. Based on our discovery experiments using purified virus, recombinant viral protein and nasopharyngeal swab samples from COVID-19 positive patients, nucleocapsid protein was selected as a target antigen. We then developed an automated antibody capture-based workflow coupled to targeted high-field asymmetric ion mobility spectrometry (FAIMS) - parallel reaction monitoring (PRM) assays on an Orbitrap Exploris 480 mass spectrometer. An ensemble machine learning-based model for determining COVID-19 positive samples was created using fragment ion intensities in the PRM data. This resulted in 97.8% sensitivity and 100% specificity with RT-PCR-based molecular testing as the gold standard. Our results demonstrate that direct detection of infectious agents from clinical samples by mass spectrometry-based assays have potential to be deployed as diagnostic assays in clinical laboratories.

## INTRODUCTION

Coronaviruses are RNA viruses and include common human coronaviruses, severe acute respiratory syndrome-CoV (SARS-CoV), Middle East respiratory syndrome-CoV (MERS-CoV) and SARS-CoV-2 (1). Although the large majority of common cold infections caused by coronaviruses do not have any significant clinical sequelae, the more recently identified coronaviruses have caused major public health outbreaks (2, 3). SARS-CoV-2 is the cause of the current COVID-19 pandemic (4, 5). Patients with COVID-19 present with upper respiratory infections that can result in a number of complications particularly in patients of advanced age or with co-morbidities. Recent reports show that in many severely ill patients it progresses to a multi-system disorder involving blood vessels (abnormal blood clotting), heart (acute cardiac injury, myocarditis and cardiac arrhythmias), kidneys (acute renal injury), liver, gut and brain (6-15).

Accurate and timely laboratory diagnosis of COVID-19 is one of the most pivotal requirements for optimal disease management and contact tracing. Historically, cell culture was the mainstay of viral antigen detection, and it still remains necessary to a minor degree, because it is the only technique capable of providing a viable isolate that can be used for further characterization of the virus. Cell culture for virus isolation also remains a useful method for growing and studying viruses in research settings especially in the characterization of novel pathogenic viruses like SARS-CoV-2 although this requires more advanced containment facilities such as biosafety level 3 (BSL-3).

The vast majority of current laboratory testing for COVID-19 is based either on detection of viral antigens, nucleic acids or on detection of virus-specific antibodies in the sera of patients. Viral antigen detection is generally based on immunoassays while viral nucleic acid detection is performed by real time quantitative RT-PCR, droplet digital PCR (ddPCR) or, less commonly, by techniques such as isothermal amplification (16). Immunoassays offer moderate to good detection sensitivity and relative rapid analytical turnaround times, but some viruses such as enteroviruses and rhinoviruses have extensive antigenic heterogeneity, which makes them less amenable to antigen detection methods. Of the molecular methods, RT-PCR and dd-PCR typically have superior detection sensitivity compared to immunoassays and less problems with viruses that show antigenic shifts, while similar turnaround times to immunoassays. Detection of virus-specific antibodies allows for diagnosis of recent symptomatic or even asymptomatic infections. IgM antibodies are the first antibody isotype that develop rapidly within days after infection, while IgG levels take a week or longer to rise. Overall, the biggest advantage of nucleic acid-based assays is that a new assay for a previously unknown virus can generally be developed and validated in a matter of a days or a few weeks after the virus is first cultured, whereas development time lines for immunoassays are substantially longer (17, 18). However, the increased demand for PCR testing worldwide has resulted in test shortages and the acute need for more available accurate diagnostic tests.

Detection of viral material using liquid chromatography-tandem mass spectrometry (LC-MS/MS) might also have sufficient sensitivity for viral antigen detection. Previous studies have suggested that detection of viral proteins in body fluids could be a rapid diagnostic method for severe acute respiratory syndrome (SARS) (19-21). Moreover, in the nearly two decades since the initial SARS outbreak, tremendous improvements have occurred in liquid chromatography as well as mass spectrometry and their applications in both research and clinical laboratories. The current generation of LC-MS-based approaches can be used for direct identification of viral proteins at relatively low concentrations and might therefore be suitable for clinical diagnostic applications. Nucleocapsid protein from SARS patients was abundant enough to be detected in clinical samples using an ELISA approach (20). Despite this, there are few, if any, studies which have focused on the detection of the viral proteins in biological samples by mass spectrometry. Targeted proteomic approaches utilizing multiple reaction monitoring or parallel reaction monitoring (MRM/PRM) are highly sensitive, allow robust detection and possess the analytical speed for clinical testing. In a pandemic situation, these approaches could supplement and complement nucleic acid based viral antigen testing. Here, we describe the development and evaluation of a mass spectrometry-based targeted assay for sensitive and accurate detection of viral antigenic peptides from nasopharyngeal swab samples of COVID-19 patients with high sensitivity and specificity.

## RESULTS AND DISCUSSION

We first sought to analyze the SARS-CoV-2 viral proteome by analyzing inactivated SARS-CoV-2 virus and recombinant viral proteins to determine which proteins and peptides are detectable. The next step was to select the most promising peptides based on spectral abundance and response intensity and to create targeted PRM methods to characterize their performance. Finally, we developed targeted assays deploying automated antibody capture-based workflow, followed by a rapid separation low-flow LC method and a PRM MS method incorporating ion mobility for selected viral peptides. In all, we tested 363 nasopharyngeal residual swab samples from patients with matched clinical molecular test results

### SARS-CoV-2 proteome

An overview of the genome organization of SARS-CoV-2 is presented in **Figure 1A**. This virus is most closely related to SARS-CoV and to MERS. The common human coronaviruses (OC43, HKU1, NL63 and L229E) are less related to SARS-CoV-2 than to each other. A sequence alignment of nucleocapsid protein in these coronaviruses is presented in **Figure S1**. A schematic of domain and functional organization of two of the major structural proteins of SARS-CoV-2, along with the location of the peptides identified in our studies is shown in **Figure 1B**.

**Figure 1.**
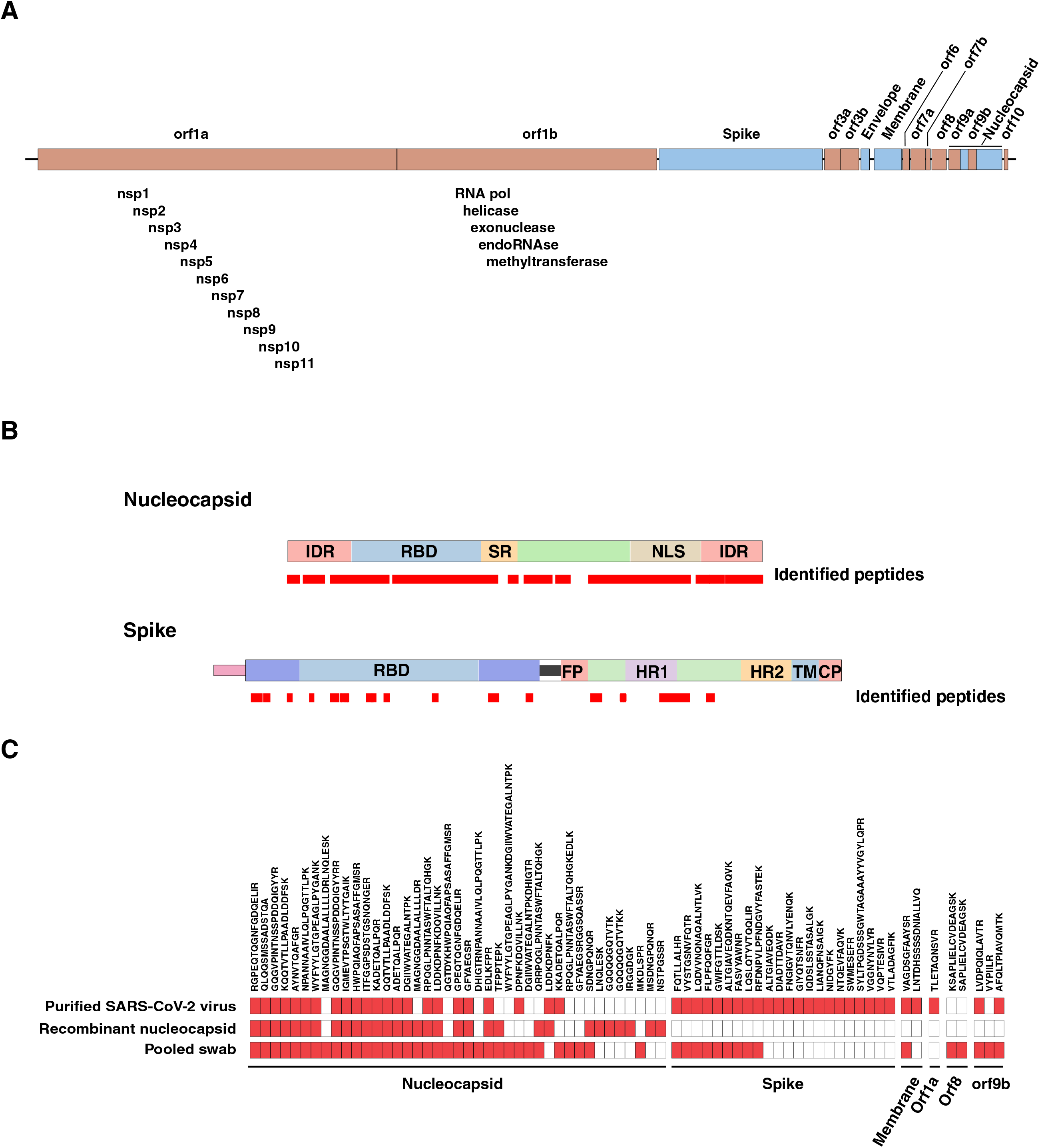
Overview of the annotated genome (A) and domain organization of key structural components: nucleocapsid and spike proteins (B) of SARS-CoV-2 along with sequence coverage obtained by peptides identified by mass spectrometry in this study. Overview of proteins and peptides from SARS-CoV-2 identified in discovery studies by bottom-up mass spectrometry (C). Red boxes represent peptides that were detected while unfilled boxes represent peptides that were not detected in the indicated samples.

### Nucleocapsid protein as a target viral antigen for assay development

To characterize the SARS-CoV-2 proteome, we analyzed purified gamma-irradiated SARS-CoV-2 virus samples (USA-WA1/2020 isolate) cultured in African green monkey *(Chlorocebus aethiops)* kidney cells (Vero E6). The inactivated virus samples were digested with trypsin and analyzed on a high resolution Orbitrap Eclipse mass spectrometer interfaced with an RSLCnano system using a 3 hr gradient. Searches against a database that included proteins from African green monkey, humans and SARS-CoV-2 plus other coronaviruses (SARS-CoV, MERS and common human coronaviruses) identified 950 proteins from monkey, 100 proteins from human (likely because of lack of a complete protein database for monkey) and 5 proteins (50 peptides) from SARS-CoV-2 including nucleocapsid (23 peptides), spike glycoprotein (22 peptides), membrane protein (2 peptides), orf1a (1 peptide) and orf9b (2 peptides) (**Table S1)**.

SARS-CoV-2 nucleocapsid is highly expressed during infection (22) and a potential target for viral detection by mass spectrometry. The nucleocapsid protein forms a supercoiled helix structure and helps package the single stranded viral RNA genome with at least 1,000 nucleocapsid molecules per virion (23, 24). Since, cells infected with SARS-CoV-2 often contain several virions, it is therefore possible to detect even relatively low viral loads in patient samples with modern LC-MS/MS instrumentation. Indeed, we consistently detected nucleocapsid-derived peptides as the most abundant peptides in our studies of the inactivated virus Trypsin digestion and analysis by untargeted LC-MS/MS of recombinant nucleocapsid protein was undertaken to identify candidate peptides for targeted detection by LC-MS/MS assays. This analysis identified 32 peptides, most of which were unique to SARS-CoV-2 (13 were indistinguishable from SARS-CoV) **(Figure S1)**. Peptides identified from SARS-CoV-2 recombinant nucleocapsid protein are summarized in **Figure 1C** and **Table S1**. To test if the peptides identified from these overexpression systems would also be identified from clinical samples, we performed LC-MS/MS-based deep proteomic profiling of pooled nasopharyngeal samples that had been confirmed by molecular test as SARS-CoV-2 positive. Database searches against human and SARS-CoV-2, SARS-CoV, MERS and other common human coronavirus proteins resulted in identification of 5,269 human proteins (50,540 peptides) and 5 proteins (55 peptides) from SARS-CoV-2 which included nucleocapsid protein (36 peptides), spike glycoprotein (3 peptides), membrane protein (1 peptide), orf9b (3 peptides) and orf8 (2 peptides), which was highly similar to the profiles previously obtained. Of the peptides that could uniquely identify SARS-CoV-2, four were chosen for targeted assay development for optimal detection (25) (**Figure 2A-D**) - all of them were unique to SARS-CoV-2 as shown by the sequence alignment with SARS-CoV, MERS and other common human coronaviruses (**Figure 2E**).

**Figure 2.**
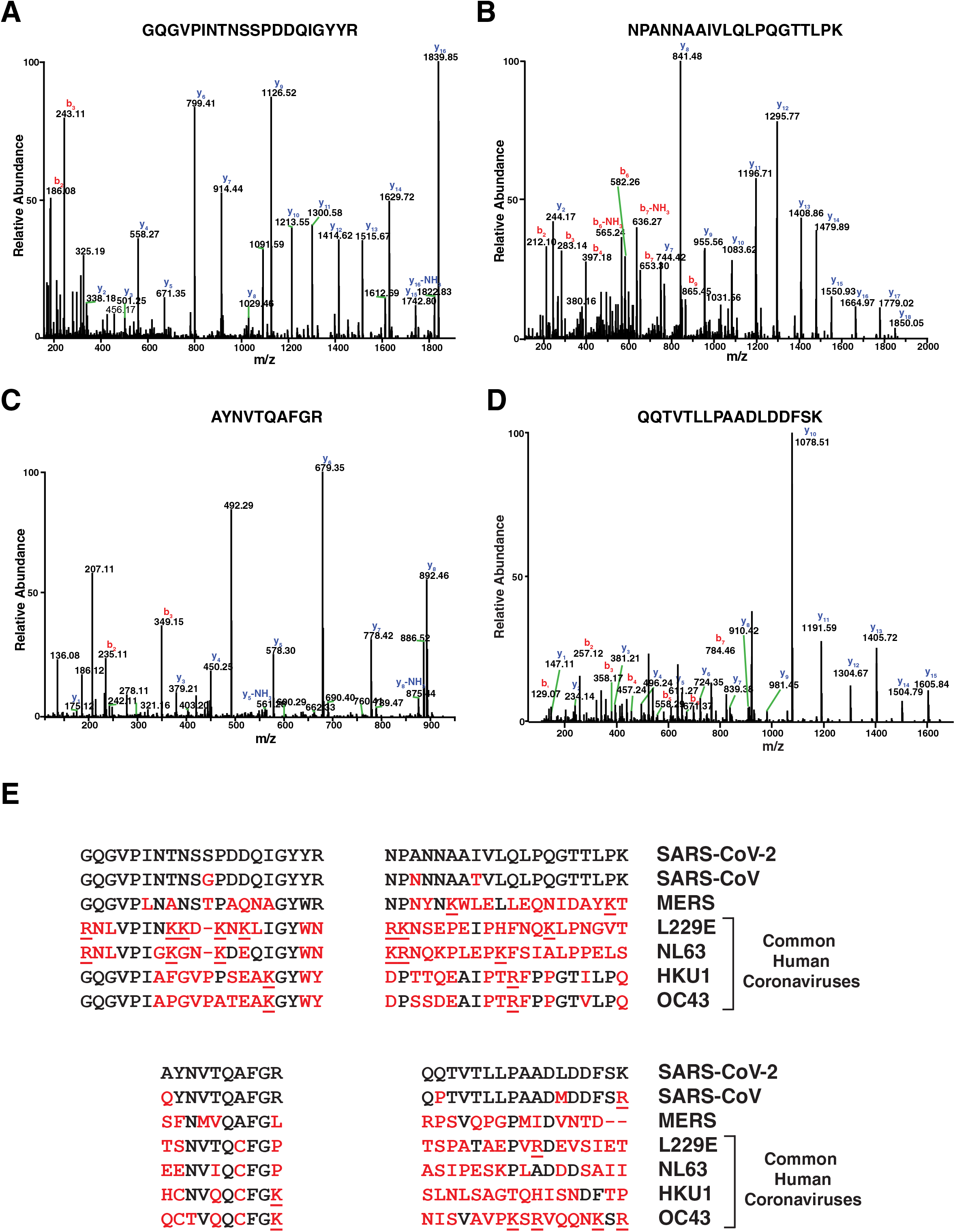
Experimental annotated MS/MS spectra of peptides identified from the nucleocapsid protein of SARS-CoV-2 positive nasopharyngeal swab specimens are shown. GQGVPINTNSSPDDQIGYYR (A), NPANNAAIVLQLPQGTTLPK (B), AYNVTQAFGR (C) and QQTVTLLPAADLDDFSK (D). Sequence alignment of the four most abundant peptides that were chosen for development of PRM assays (E) - GQGVPINTNSSPDDQIGYYR (69-88), NPANNAAIVLQLPQGTTLPK (150-169), AYNVTQAFGR (267-276) and QQTVTLLPAADLDDFSK (389-405) derived from SARS-CoV-2 nucleocapsid protein across related coronaviruses (SARS-CoV, MERS and common human coronaviruses: L229E, NL63, HKU1 and OC43). Amino acid mismatches are indicated in red while tryptic cleavage sites (R/K) are underlined.

### Optimization of pre-analytical variables

Based on our discovery experiments, we decided upon nucleocapsid protein as our target antigen for assay development. We optimized several pre-analytical parameters including protein extraction, various antibodies for capture of nucleocapsid protein and the use of ion mobility.

#### A. Protein extraction optimization

To maximize protein yield from nasopharyngeal swab samples, we tested several protein extraction methods including methanol precipitation, 2% SDS, 0.002% Z316 zwitterionic detergent and RIPA buffer. The processed samples were immunopurified and MS/MS fragment intensities for AYNVTQAFGR and QQTVTLLPAADLDDFSK peptides were compared. From these experiments, 0.002% Z316 was selected for protein extraction and subsequent capture using antibody.

#### B. Direct digest and immunopurification

Next, we tested if we could detect the nucleocapsid protein directly after proteolytic digest or if enrichment using an anti-nucleocapsid antibody prior to digestion was required for detection. For this comparison, a monoclonal antibody (Sino Biological, Wayne PA, Cat# 40143-R001) was biotinylated and immobilized to streptavidin mass spectrometry immunoassay (MSIA) tips for capture using an automated liquid handler. In all, 89 patient samples were analyzed with each method (69 positive and 20 negative as tested by RT-PCR). To determine if antibody enrichment resulted in an increase in sensitivity, a cutoff was established at 3 standard deviations above the average peak height observed for AYNVTQAFGR and QQTVTLLPAADLDDFSK in the 20 RT-PCR negative samples. Peptide signals above the cutoff threshold in RT-PCR positive samples were then counted. Antibody enrichment resulted in 14 and 24 additional positive samples which were above the cutoff for AYNVTQAFGR and QQTVTLLPAADLDDFSK, respectively. The additional samples above the cutoff in antibody enriched samples had higher Ct values (i.e. lower viral loads) as compared to samples above the cutoff for both the methods, indicating that immunoaffinity enrichment results in a greater sensitivity relative to direct digest (**Table S2)**.

#### C. Antibody enrichment optimization

To improve the efficiency of nucleocapsid capture, 17 different antibodies against the nucleocapsid protein were evaluated (**Table S3**). Four antibodies which displayed the best performance characteristics were compared side by side using pooled patient samples with high, medium or low viral loads (based on Ct values from real time quantitative RT-PCR-based molecular testing). This comparison identified two monoclonal antibodies with suitable performance from which we selected one for further development of the assay.

#### D. Improved sensitivity with ion mobility

In discovery LC-MS/MS analysis of eluates after nucleocapsid enrichment from nasopharyngeal samples, we detected >2,000 peptides. This poses unique challenges especially if short chromatographic separation is desired for targeted detection owing to interference from the highly complex matrix. Orthogonal gas phase separation such as ion mobility for separation of charged ions based on their size could potentially provide additional separation that might be beneficial. A front-end high-field asymmetric waveform ion mobility spectrometry (FAIMS) source typically filters ions entering into the ion transfer capillary by reducing background chemical noise resulting in increased sensitivity and robustness (26, 27)To determine if FAIMS could increase specificity for the viral peptides, we optimized compensation voltages for the peptides of interest from nucleocapsid protein. Those that resulted in the highest signal intensities were chosen to build the targeted method **(Table S4)**. We analyzed nasopharyngeal swab samples with and without FAIMS and observed significantly improved signal-to-noise (S/N) ratio with FAIMS. **Figure S2A** shows the PRM signal of AYNVTQAFGR peptide from a representative nasopharyngeal swab sample (relatively high viral load; Ct value of 20) where the S/N with FAIMS was 965 as compared to 67 without FAIMS. **Figure S2B** shows the PRM signal for AYNVTQAFGR peptide in another representative nasopharyngeal swab sample with a low viral load (Ct value of 30) that could not be detected without FAIMS. Based on these data, we incorporated FAIMS into our assay workflow.

### High-throughput assays for detection of SARS-CoV-2

After optimization of protein extraction, capture and LC-MS/MS acquisition parameters, we selected two peptides for the final targeted assay. To maintain optimal performance, the sample preparation and chromatography conditions were further refined. The sample preparation workflow was automated using Versette liquid handler which minimized overall time and experimental variation during sample processing. The throughput and robustness of the sample analysis was achieved by coupling Evosep One with Exploris 480 mass spectrometer, allowing analysis of 100 samples per day without sacrificing sensitivity.

Synthetic isotopically labeled heavy peptides were used for optimization of LC-MS/MS parameters including collision energy, retention time and relative intensity of fragment ions. **Table 1** shows details of peptides including sequence, precursor m/z, charge, retention time and FAIMS compensation voltages. This optimized assay was used to analyze 363 residual nasopharyngeal swab samples previously tested by quantitative real time RT-PCR. **Figure 3** shows a schematic of optimized workflow for the diagnosis of SARS-CoV-2 using FAIMS-PRM targeted mass spectrometry.

**Figure 3.**
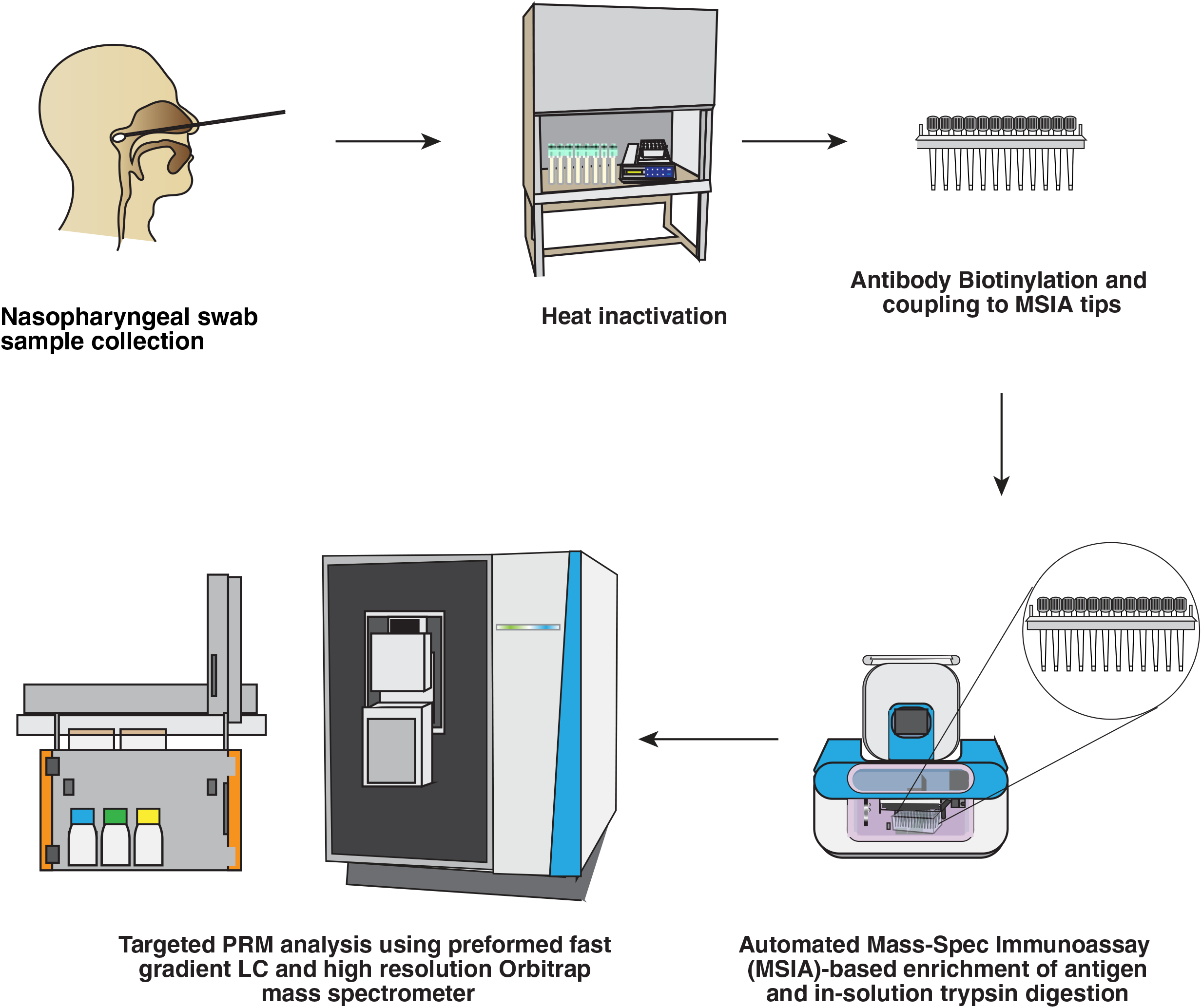
A schematic of FAIMS-PRM targeted assay for the detection of diagnosis of SARS-CoV-2. Heat-inactivated nasopharyngeal swab samples were immunopurified with anti-nucleocapsid antibody coated on MSIA D.A.R.T.s™ tips using Versette automated liquid handler, in-solution trypsin digested. Targeted LC-MS/MS analysis was carried out on Exploris 480 mass spectrometer interfaced with rapid chromatography system and a FAIMS Pro interface.

**Table 1:** Peptides from nucleocapsid protein used for targeted FAIMS-PRM analysis for the detection of SARS-CoV-2 in nasopharyngeal swab samples. The table includes peptide sequence, precursor *m/z*, precursor charge, retention time (start and end) in minutes and optimized compensation voltage (CV).

| Peptide | m/z | Charge State | Start (min) | End (min) | FAIMS Pro CV |
| --- | --- | --- | --- | --- | --- |
| AYNVTQAFGR | 563.7856 | 2 | 2.5 | 3.5 | -40 |
| AYNVTQ**A**(+3Da)FGR(+10Da) (Heavy IS) | 570.2948 | 2 | 2.5 | 3.5 | -40 |
| QQTVTLLPAADLDDFSK | 931.4807 | 2 | 3.5 | 5.3 | -30 |
| QQTVTLLPAAD**L**(+7Da)DDFS**K**(+8Da) (Heavy IS) | 938.9964 | 2 | 3.5 | 5.3 | -30 |

We analyzed 187 nasopharyngeal swab samples (116 positive and 71 negative) using the optimized FAIMS-PRM workflow and used supervised machine learning to select the optimal fragments and determine their weights for maximizing the performance of the targeted mass spectrometry assay. Using this as a training dataset, we employed an ensemble-based machine learning approach encoded in Super Learner as described previously (28). This method was configured to use a generalized linear model via penalized maximum likelihood (glmNET), generalized linear model (GLM) and random forest (RF) model; all configured to use binomial distribution. A 10-fold cross-validation with a goal to maximize the AUC and to prevent overfitting was instituted during the learning process. An optimal weighted average of the trained models (i.e glmNET, GLM and RF) was computed and considered as a final composite model to evaluate performance on an independent validation dataset. This ensemble method incorporated a total of 17 fragments, resulting in an AUC of 0.9956 on the training set. Based on the probability distribution of negative and positive samples in the training set, we chose a probability of ≥0.6 as threshold for calling a positive sample **(Figure 4A and 4B)**. We applied this composite model to an independent validation dataset of 176 samples (88 positive and 88 negative samples) using the exact same optimized FAIMS-PRM workflow. This resulted in achieving a sensitivity of 97.8% and specificity of 100% with an AUC=1.0. These results show that our assay for the detection of SARS-CoV-2 antigen performed with a very high sensitivity - and specificity, especially compared to other reported rapid antigen lateral flow assays (29).

**Figure 4.**
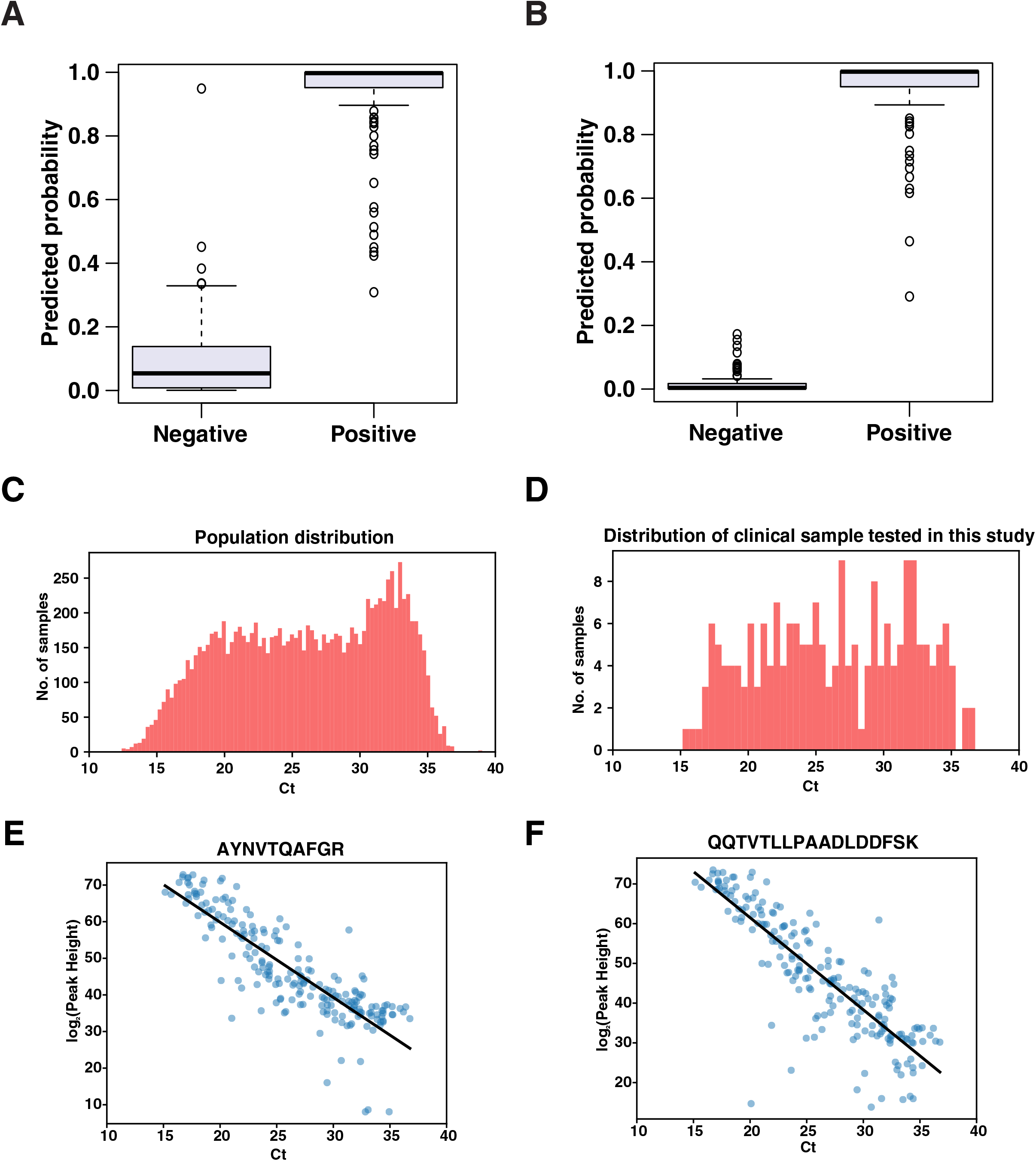
Box plot showing predicted probability for training dataset (A) and validation dataset (B). Population distribution of nasopharyngeal swab samples tested by RT-PCR assay at Mayo Clinic (C), Distribution of clinical samples tested using targeted FAIMS-PRM method in this study (D), Targeted FAIMS-PRM fragment ion intensity against Ct values of nasopharyngeal swab samples for AYNVTQAFGR (E) and QQTVTLLPAADLDDFSK (F) peptides.

### Assay performance as compared to RT-PCR tests

Although RT-PCR remains the well-established gold standard for viral pathogen detection, the FAIMS-PRM workflow displays very high sensitivity and specificity. To ensure that samples used for training and validation of the model reflect the distribution observed in a large population, we mimicked the distribution of Ct values for 11,575 SARS-CoV-2 positive patients tested at Mayo Clinic (**Figure 4C**) in the training and test set samples (**Figure 4D**). Ct values obtained from the large population approximate a normal distribution (mean = 26.5, IQR = 21.1 - 31.3) and agree well with the training and validation samples (mean = 26.5, IQR = 21.8 - 31.4). While naïve, uncalibrated Ct values are not themselves quantitative they can serve as an estimate of relative target abundance. Peptide peaks heights derived from mass spectrometry analysis also provide semi quantitative information. To determine the feasibility of using peak heights as an estimate of viral abundance Ct values were correlated to peak heights using the training and validation data sets (**Figure 4E** and **4F**). Indeed, both peptides display good correlation with Ct values (QQTVTLLPAADLDDFSK r^2^ = 0.73, AYNVTQAFGR r = 0.73) indicating that viral peptide peak height may additionally serve as an estimate of viral abundance.

## Conclusions

In this study, we report development of a mass spectrometry-based targeted assay that permits direct detection of viral antigens from clinical specimens. A key advantage of this targeted method is the high specificity and comparable sensitivity to the gold standard RT-PCR method for the diagnosis of SARS-CoV-2. Definitive identification by this method can also serve as a reference method for other antigen testing assays such lateral flow assays, many of which lack sensitivity. In addition, the method can be applied to other sample types (e.g. urine, saliva) and can be adapted for detection of novel variants of SARS-CoV-2 as they appear. Finally, our method with rapid chromatographic separation followed by highly sensitive mass spectrometry detection provides a throughput of 100 samples per day and can be easily deployed at routine clinical laboratories with mass spectrometry instrumentation.

## Data Availability

The mass spectrometry proteomics data have been deposited to the ProteomeXchange Consortium via the PRIDE partner repository with the dataset identifier PXD020644.
Reviewer account details:
Username:
Password: J8setKN6

## Acknowledgments

The following reagent was deposited by the Centers for Disease Control and Prevention and obtained through BEI Resources, NIAID, NIH: SARS-Related Coronavirus 2, Isolate USA-WA1/2020, Gamma-Irradiated, NR-52287. This study was supported by ‘DBT/Wellcome Trust India Alliance’ Margdarshi Fellowship grant IA/M/15/1/502023 awarded to AP.

## Data Availability

The mass spectrometry proteomics data have been deposited to the ProteomeXchange Consortium via the PRIDE partner repository with the dataset identifier PXD020644.

Reviewer account details:

Username: revi ewer9 Password: J8setKN6

## MATERIALS AND METHODS

### COVID-19 specimen collection and handling

All samples were collected after informed consent and approval by the institutional review board. All clinical samples were de-identified prior to analysis.

### Chemicals and reagents

The following reagents were deposited by the Centers for Disease Control and Prevention and obtained through BEI Resources, NIAID, NIH: SARS-Related Coronavirus 2, Isolate USA-WA1/2020, gamma-irradiated, NR-52287. Tris (2-carboxyethyl)phosphine hydrochloride (TCEP) and iodoacetamide were purchased from Sigma (St. Louis, MO, USA). Phosphate buffered saline (PBS) was purchased from Bio-Rad (Hercules, CA, USA). Trifluoroacetic acid (TFA) was purchased from Thermo Fisher Scientific (Waltham, MA, USA). Zwittergent Z3-16 was purchased from CalBiochem (EMD Millipore, Billerica, MA, USA). Dynabeads M-280 Streptavidin was purchased from Invitrogen (Carlsbad, CA, USA). Trypsin/Lys-C Mix and Rapid Digestion kit was purchased from Promega (Madison, WI, USA). Recombinant SARS-CoV-2 nucleocapsid protein (97-077) was purchased from ProSci (Fort Collins, CO).

### Synthesis of isotopically labeled heavy peptide standards

All SARS-CoV-2-derived peptides were synthesized using standard FMOC chemistry on a Liberty Blue (CEM Corp. Matthews, NC) peptide synthesizer with methods suggested by the manufacturer. Starting resin was either arginine (pbf) ^13^C_6_, ^15^N_4_ 2Cl-trt resin or lysine (boc) ^13^C_6_ ^15^N_2_ 2Cl-trt resin (Cambridge Isotope Laboratories, Tewksbury, MA), depending on the sequence. A second stable isotope labeled amino acid was incorporated into the sequence using Fmoc-Leucine-OH ^13^C_6_ ^15^N, Fmoc-Glycine-OH ^13^C_2_ ^15^N, or Fmoc-Alanine-OH ^13^C_3_ (IsoTec, Inc., Miamisburg, OH) as dictated by the sequence. Peptides were cleaved using the CEM Razor cleavage module heated to 42°C for 40 min. Cleavage cocktail was trifluoroacetic acid, water, triisopropyl silane and 3,6-Dioxa-1,8-octanedithiol (92.5/2.5/2.5/2.5 v/v/v/v). Peptides were precipitated in cold methyl-t-butyl ether, washed and dried for purification. Purification was achieved by reversed-phase HPLC using a Phenomenex Jupiter C_18_ column, 250 x 21.2 mm, using a water/acetonitrile buffer system. Peptide purity and integrity were confirmed using an Agilent InfinityLab II LC/ MSD (Agilent Technologies, Santa Clara, CA) system.

### Processing of nasopharyngeal swab samples and in-solution trypsin digestion

Nasopharyngeal swab samples were collected in PBS and digested directly in-solution. 100 μl of sample was diluted three-fold with rapid digest buffer (Promega, Madison WI) and 1.5 μg of trypsin was added. The samples were placed on a thermomixer (Thermo Scientific, Waltham, MA) at 70°C for 1 hr with rotation at 1,150 rpm for trypsin digestion. To terminate the digestion, the samples were acidified to 0.2% TFA. The samples were desalted using C_18_ solid phase extraction spin columns (Glygen, Columbia, MD), dried down and reconstituted in 20 μl of 0.2% formic acid. Recombinant proteins and inactivated purified SARS-CoV-2 were digested using the method described above for the swab samples with the following modifications: protein content of the purified SARS-CoV-2 was estimated using a BCA assay (Pierce Waltham, MA) and trypsin was added to all samples at a 1:10 enzyme to substrate ratio. Following digestion, disulfide bonds were reduced and cysteines were alkylated simultaneously with the addition of 5 mM TCEP and 5 mM IAA. Samples were incubated in dark for 30 min at room temperature prior to sample acidification and cleanup.

### Untargeted LC-MS/MS experiments

LC-MS/MS analysis for untargeted discovery proteomics experiments was carried out using an Ultimate 3000 RSLCnano system (Thermo Scientific, San Jose, CA) connected to an Orbitrap Eclipse mass spectrometer (Thermo Scientific, San Jose, CA). The peptides were loaded onto a trap column (PepMap C_18_ 2 cm x 100 μm, 100 Å) at a flow rate of 20 μl/min using 0.1% formic acid and separated on an analytical column (EasySpray 50 cm x 75 μm, C_18_ 1.9 μm, 100 Å, Thermo Scientific, San Jose, CA) with a flow rate of 300 nl/min with a linear gradient of 5 to 40% solvent B (100% ACN, 0.1% formic acid) over a 40 min gradient. Both precursor and fragment ions were acquired in the Orbitrap mass analyzer. Precursor ions were acquired in *m/z* range of 350-1700 with a resolution of 120,000 (at *m/z* 200). Precursor fragmentation was carried out using higher-energy collisional dissociation (HCD) method using normalized collision energy (NCE) of 28. The fragment ions were acquired at a resolution of 30,000 (at *m/z* 200). The scans were arranged in top-speed method with 3 seconds cycle time between MS and MS/MS. Ion transfer capillary voltage was maintained at 2.5 kV. For internal mass calibration, lock mass option was enabled with polysiloxane ion *(m/z*, 445.120025) from ambient air.

### Anti-nucleocapsid antibody-based enrichment of nucleocapsid protein and in-solution trypsin digestion

In order to improve the sensitivity of the detection, we evaluated a number of antibodies as shown in **Table S3**. Briefly, antibody was biotinylated using biotinylation kit (ThermoFisher Scientific, San Jose, CA) as per manufacturer’s instructions. Biotinylated antibody (1 μg) was coated on streptavidin MSIA tips (Catalog#991STR11, ThermoFisher Scientific, San Jose, CA) in 0.1% BSA containing 1X PBS on the Versette automated liquid handler (ThermoFisher Scientific, San Jose, CA). Nasopharyngeal swab samples (750 μl) were mixed with zwitterion Z316 at final concentration of 0.002% in 96 well plate and were inactivated at 70°C for 30 minutes. Inactivated samples were subjected to enrichment using mass spectrometry immunoassay (MSIA)-based enrichment using biotinylated antibody, washed two times with 200 μl 1X PBS and eluted in 100 μl of 50% ACN/0.002% Z316 in 0.1% TFA. Sample eluent was mixed with 300 μl of rapid trypsin digestion buffer (Promega Corporation, Madison, WI) and subjected to in-solution trypsin digestion (Gold Trypsin, Promega Corporation, Madison, WI) at 70°C for 1 hour on a shaker incubator. The digest was acidified using TFA to a final concentration of 1% TFA. The acidified digests were spiked-in with synthetic isotope labelled heavy peptides and the samples were loaded on EvoTips as per manufacturer’s instructions. Briefly, the C_18_ EvoTips were activated using 20 μl of 100% acetonitrile followed by equilibration with 20 μl of 0.1% formic acid in water. Activation and equilibration was carried out at 700 x g for 1 minute. The sample was loaded at 500 x g for 5 minute followed by washing using 0.1% formic acid once. At last the tips were loaded with 100 μl of 0.1% formic acid and processed for targeted analysis.

### Targeted Parallel Reaction Monitoring (PRM) analysis

Parallel reaction monitoring (PRM) analysis was performed on an Exploris 480 mass spectrometer equipped with FAIMS Pro ion source (Thermo Scientific, Bremen, Germany) and interfaced with a preformed gradient LC system (EvoSep One, EvoSep Inc., Odense, Denmark). FAIMS Compensation voltages were optimized for peptides from nucleocapsid protein; AYNVTQAFGR and QQTVTLLPAADLDDFSK. Individual positive and negative swab samples were processed for antibody capture and trypsin digestion as described above. Peptides were eluted at a flow rate of 2 μl / minute into a pre-formed gradient using the Evosep One LC system connected to the Exploris 480 mass spectrometer equipped with FAIMS Pro ion source. Peptide separation was carried out using a 4 cm analytical column (Dr. Maisch C_18_AQ, 1.9 μm, 150 μm x 4 cm) with a 5.6 min gradient. Data acquisition parameters included MS1 scan from 560-100 m/z at resolution of 60,000 followed by retention time scheduled PRM analysis of AYNVTQAFGR and QQTVTLLPAADLDDFSK peptides and corresponding double SIL heavy peptides. The PRM parameters included: Orbitrap resolution of 60,000, AGC target value of 5 x 10^4^, injection time of 118 ms, isolation window of *m/z* 1 and HCD normalized collision energy of 27. Table 1 shows targeted inclusion list with retention time scheduled PRM scans for light and heavy versions of AYNVTQAFGR and QQTVTLLPAADLDDFSK with corresponding FAIMS compensation voltages (CV).

### Mass spectrometry data analysis of untargeted LC-MS/MS data

The raw mass spectrometry data were searched using Andromeda in MaxQuant software suite (version 1.6.7.0) (30) against a combined protein database of SARS-CoV-2 proteins, SARS-CoV proteins, common coronaviruses (OC43, HKU1, NL63 and L229E) and UniProt human protein database, African green monkey *(Chlorocebus aethiops)* database (in case of irradiated virus MS data) including common MS contaminants. The search parameters included a maximum of two missed cleavages; carbamidomethylation at cysteine as a fixed modification for samples that were reduced and alkylated; N-terminal acetylation and oxidation at methionine as variable modifications. Precursor tolerance was set to 10 ppm and MS/MS tolerance to ±0.02 Da. False discovery rate was set to 1% at the peptide-spectrum matches (PSMs), peptide and protein levels.

### Data analysis of targeted LC-MS/MS data

The PRM data were processed using the Skyline software package (31). PRM data were assessed for (i) peak symmetry and baseline; (ii) consistency of retention times and (iii) retention time consistency across transitions of the same analyte. In addition, peak integration of extracted ion chromatogram of all plausible fragments of light and heavy precursor was adjusted manually to avoid potential interferences. The fragment ion intensities were exported from skyline and (natural) log transformed. A supervised machine learning method was used to select the optimal fragments and determine their weights for maximizing the performance of the targeted mass spectrometry assay. All computations were performed in R (version 4.0.1). For this, we utilized an ensemble-based machine learning approach encoded in the Super Learner as described previously. This method was configured to use a generalized linear model via penalized maximum likelihood (glmNET), generalized linear model (glm) and random forest model; all configured to use binomial distribution. A 10-fold cross-validation with a goal to maximize the AUC was instituted during the learning process. An optimal weighted average of the different trained models was computed and considered as final model for an independent validation.

### SUPPLEMENTARY FIGURES

**Figure S1.**
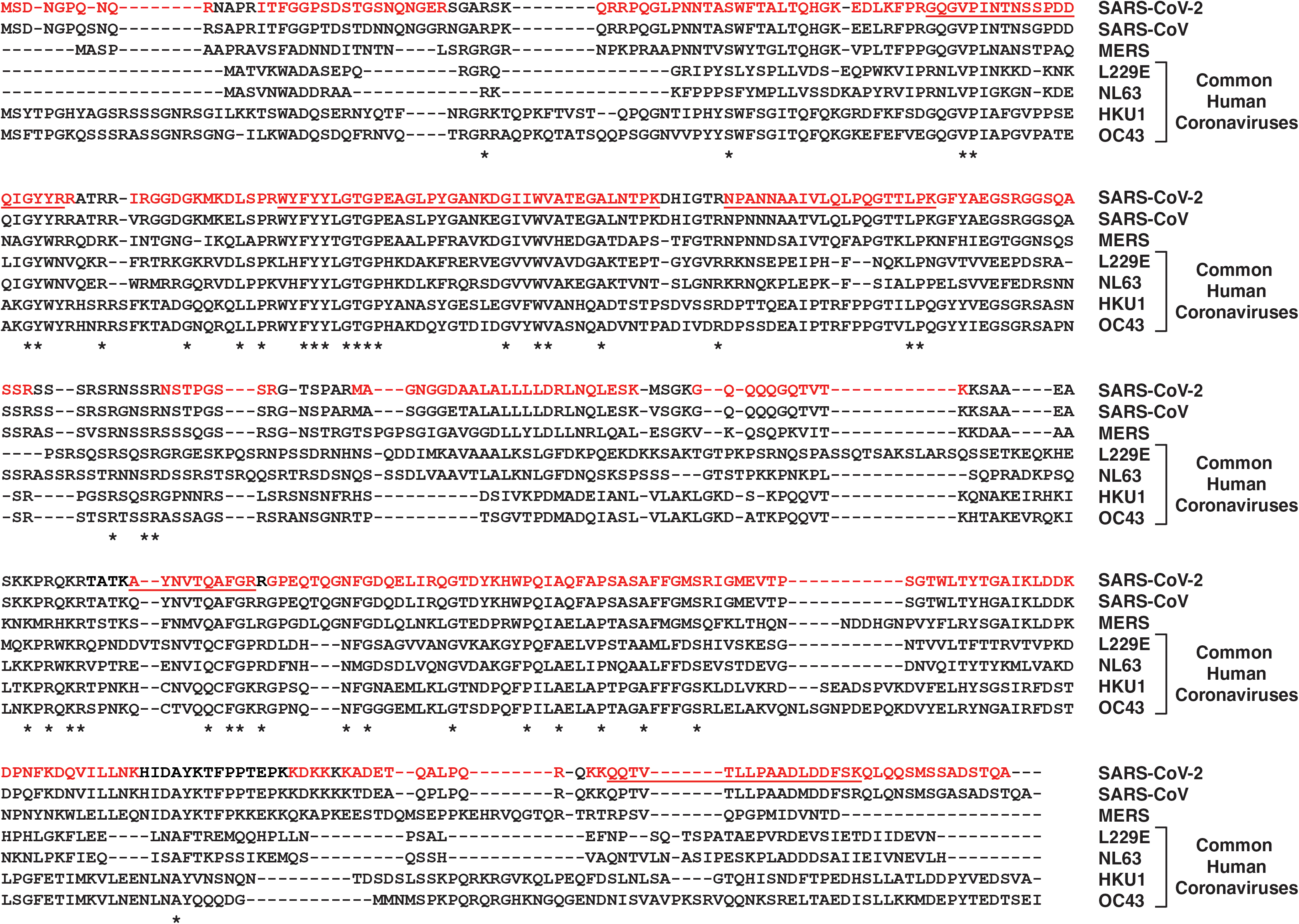
Sequence alignment of SARS-CoV-2 nucleocapsid protein against related coronaviruses: SARS-CoV, MERS and common human coronaviruses (L229E, NL63, HKU1 and OC43). Sequence in red indicates the regions that were identified by mass spectrometry while the underlined sequences were selected for targeted PRM analysis.

**Figure S2.**
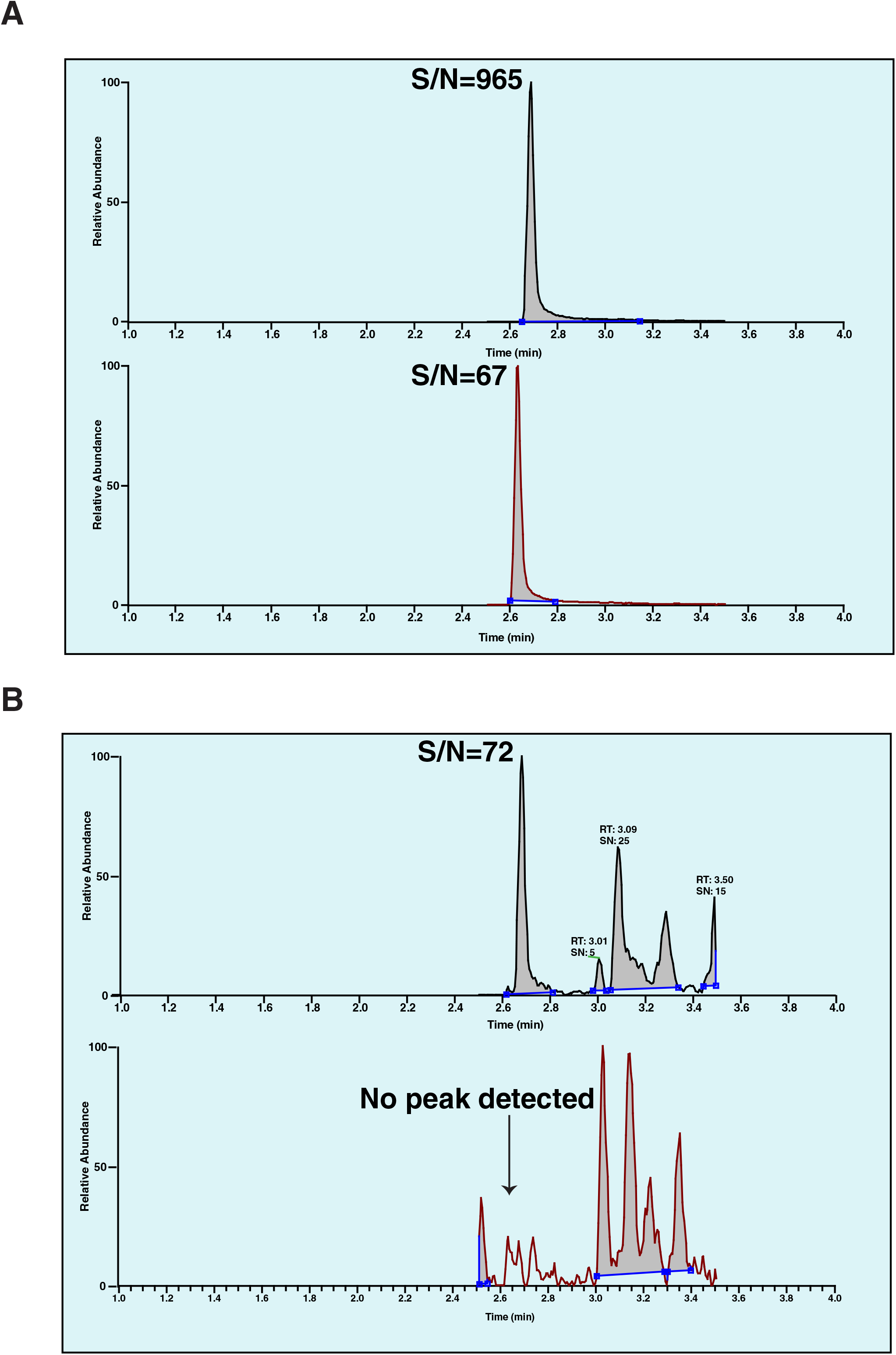
Extracted ion chromatogram (EIC) for the top three fragments (m/z 678.36, 778.43, 892.47) of AYNVTQAFGR peptide *(m/z* 563.78) in nasopharyngeal swab IP samples with Ct value of 20 (A) and Ct value of 30 (B). Top panels in both (A) and (B) are with FAIMS while bottom panels are without FAIMS.

### SUPPLEMENTARY TABLES

Table S1: List of all proteins and peptide identified by mass spectrometry across various sample types analyzed in this study.

Table S2: Fragment ion intensities of direct digest and immunopurified samples analyzed by targeted PRM experiments.

Table S3: List of all antibodies used for evaluation for enrichment and detection of nucleocapsid and/or spike glycoprotein from SARS-CoV-2

Table S4: Fragment ion intensities of nasopharyngeal swab IP samples analyzed by FAIMS and noFAIMS targeted PRM method.

Table S5: Fragment ion intensities of nasopharyngeal swab IP samples analyzed used for machine learning training and validation.

